# The Impact of U.S. County-Level Factors on COVID-19 Morbidity and Mortality

**DOI:** 10.1101/2021.01.19.21250092

**Authors:** Nevo Itzhak, Tomer Shahar, Robert Moskovich, Yuval Shahar

## Abstract

**Background:** The effect of socioeconomic factors, ethnicity, and other variables, on the frequency of COVID-19 cases [morbidity] and induced deaths [mortality] at *sub-population*, rather than at *individual* levels, is only partially understood.

**Objective:** To determine which county-level features best predict COVID-19 morbidity and mortality for a given county in the U.S.

**Design:** A Machine-Learning model that predicts COVID-19 mortality and morbidity using county-level features, followed by a SHAP-values-based importance analysis of the predictive features.

**Setting:** Publicly available data from various American government and news websites.

**Participants:** 3,071 U.S. counties, from which 53 county-level features, as well as morbidity and mortality numbers, were collected.

**Measurements:** For each county: Ethnicity, socioeconomic factors, educational attainment, mask usage, population density, age distribution, COVID-19 morbidity and mortality, air quality indicators, presidential election results, ICU beds.

**Results:** A Random Forest classifier produced an AUROC of 0.863 for morbidity prediction and an AUROC of 0.812 for mortality prediction. A SHAP-values-based analysis indicated that poverty rate, obesity rate, mean commute time to work, and proportion of people that wear masks significantly affected morbidity rates, while ethnicity, median income, poverty rate, and education levels, heavily influenced mortality rates. The correlation between several of these factors and COVID-19 morbidity and mortality, from 4/2020 to 11/2020 shifted, probably due to COVID-19 being initially associated with more urbanized areas, then with less urbanized ones.

**Limitations:** Data are still coming in.

**Conclusions:** Ethnicity, education, and economic disparity measures are major factors in predicting the COVID-19 mortality rate in a county. Between-counties low-variance factors (e.g., age), are not meaningful predictors.

Differing correlations can be explained by the COVID-19 spread from metropolitan to less metropolitan areas.

**Primary Funding Source:** None.

## 1 Introduction

Since December 2019, the novel Coronavirus disease 2019 (COVID-19) has spread rapidly around the globe, infecting millions, leading to severe illness, hospitalization, admission to intensive care units (ICUs) [1], and death. In the U.S., more than 20 million people have been infected and some 400,000 died from COVID-19.

Individual-patient predictors include age, gender, and ethnicity [2,3,4]. However, both infection and death rates vary highly among different countries and regions and are not easy to predict [5].

It is important to comprehend the population-level factors affecting morbidity and mortality due to COVID-19, to better appreciate the effect of various interventions, prepare for expected morbidity and mortality in different regions, and apply the preventive and therapeutic measures most appropriate for each site (e.g., vaccination).

Population density does not seem to be linked with COVID-19 morbidity or mortality in the U.S. [6] and China [7], but has been linked in countries such as India [8]. A few studies have found that prolonged exposure to poor air quality may lead to more COVID-19 deaths [9]. Other researchers, such as Chang et al. [10], demonstrated that a small minority of *superspreader* crowded points of interest, such as restaurants and grocery stores, account for a large majority of the infections. Higher infection rates among disadvantaged racial and socioeconomic groups seemed to result solely from differences in mobility, due to lack of ability to work from home.

The spread of COVID-19 at the county level for the U.S. was also studied by assessing the counties’ vulnerability [11,12] or by assessing the effect of county-level features [13,14,15]. Statistical analysis was the popular approach [13,15,11]; however, some studies used a machine learning approach [14,12]. Counties with a larger percentage of racial and ethnic minorities were affected the most [14,15,13]. Millett et al. [13] found that counties who had a large percentage (greater than 13 percent) of African Americans accounted for more than half of the cases and deaths nationally. Cahill et al. [11] found that counties with a lower *Case Fatality Rate* (CFR) had a greater proportion of the population reporting having two or more races. However, no significant differences were found between High and Low CFR counties with respect to mean income or poverty rate.

Most studies used a *subset* of the features we are analyzing in the current study; none contain *all* of the ethnicity, socioeconomic, epidemiological factors, population density, and population age features, as well as additional features, such as mask usage, and even presidential election results.

Two studies comparable to ours also adopted machine learning approaches. Tiwari et al. [12] built a machine learning model to measure county-level vulnerability, then overlaid the vulnerability map on county-level features, such as on the racial minority population percentage data. Paul et al. [14], like us, examined factors that contribute to COVID-19 prevalence or death rate using machine learning, and then an approach inspired by game theory to calculate the features’ importance. Unlike us, these authors solved the task using *regression* instead of *classification*.

Our study’s goals were:

1. Determine, using a Machine-Learning approach, coupled with a method inspired by Game Theory, the relative impact of a large number of static factors, such as ethnicity, socioeconomic status, and self-reported behavior, on sufficiently accurate machine-learning models that we have built, for prediction of COVID-19 (*morbidity*) and (*mortality*) for any county in the U.S. at any time;
2. Determine how this impact varies over *time*, and the trends characterizing the varying importance (Note: We did not try to *predict* any *future* COVID-19 morbidity or mortality rates. Our approach is purely classification based.)

## 2 METHODS

The U.S. is comprised of 3,243 counties; we have gathered COVID-19 related data for 3,071 of these counties.

Specifically, we gathered ethnicity, socioeconomic (e.g., income, mode of transportation), educational attainment, epidemiological factors, ICU bed availability, mask usage, presidential election results, population density and age and gender distribution across multiple age groups.

The data were extracted from multiple sources. The full list of data types and of their sources appears in the *Supplementary Materials*.

Our goal was to assess the relative importance of the features and examine their impact on the classification of each county’s morbidity and mortality rates. To do this, we trained two separate classification models; one for predicting the morbidity rate, and one for predicting the mortality rate, in any given county. We treated the task as a *binary classification* problem. A county in the top quartile of morbidity rates (5.8 percent or higher) was labeled as “High Morbidity”, otherwise as “Low Morbidity”. Similarly, a county in the top quartile of mortality rates (0.1 percent or higher) received a label of “High Mortality”, otherwise “Low Mortality”.

Morbidity and mortality rates across the U.S. can be viewed in the supplementary materials.

Both COVID-19 morbidity and mortality classification models were induced using a Random Forest algorithm. We used hyper-parameter optimization to fit the best parameters to our model, using a ten-fold cross-validation.

The details of inducing the classification model are provided in the Supplementary Materials.

Once the model converged, and we confirmed that it achieved a sufficiently high accuracy score, the *SHapley Additive exPlanations* (SHAP) [16] approach, inspired by an established Game Theory result that determines the importance of different players in various coalitions, was used to determine the absolute impact (and direction of influence) of different variables on the classification models’ output, through a ten-fold cross-validation.

The details of building the SHAP model are provided in the Supplementary Materials.

## 3 RESULTS

After removing highly correlated features such as the percentage of females and males, median male age and median total age, and voting statistics for the democratic and liberal parties, the dataset used, representing the integrated data of 3,071 counties, included 53 features. The Random Forest morbidity and mortality classification models we have built were based on these features.

The model that predicted the morbidity level resulted in an *Area Under the ROC curve* (AUROC) of 0.863 and an *area Under the Precision-Recall Curve* (AUPRC) of 0.697.(See Supplementary Materials for evaluation metrics’ details).

The model that predicted the mortality rates resulted in an AUROC of 0.812 and an AUPRC of 0.629.

Thus, we considered both models as sufficiently accurate for the impact weights to be meaningful.

Figure 1 presents the SHAP summary output, which shows the impact of the features on the COVID-19 morbidity (Fig. 1.a) and mortality (Fig. 1.b) models that were induced, on all of the individual instances of all counties on which the model was tested. The figure sorts the features in a top-down fashion by the absolute sum of the SHAP value magnitudes over all samples, and uses the SHAP values to show the distribution of the impacts each feature has on each model output. Only the top twenty features are presented; the rest were less impactful.

**Fig. 1.**
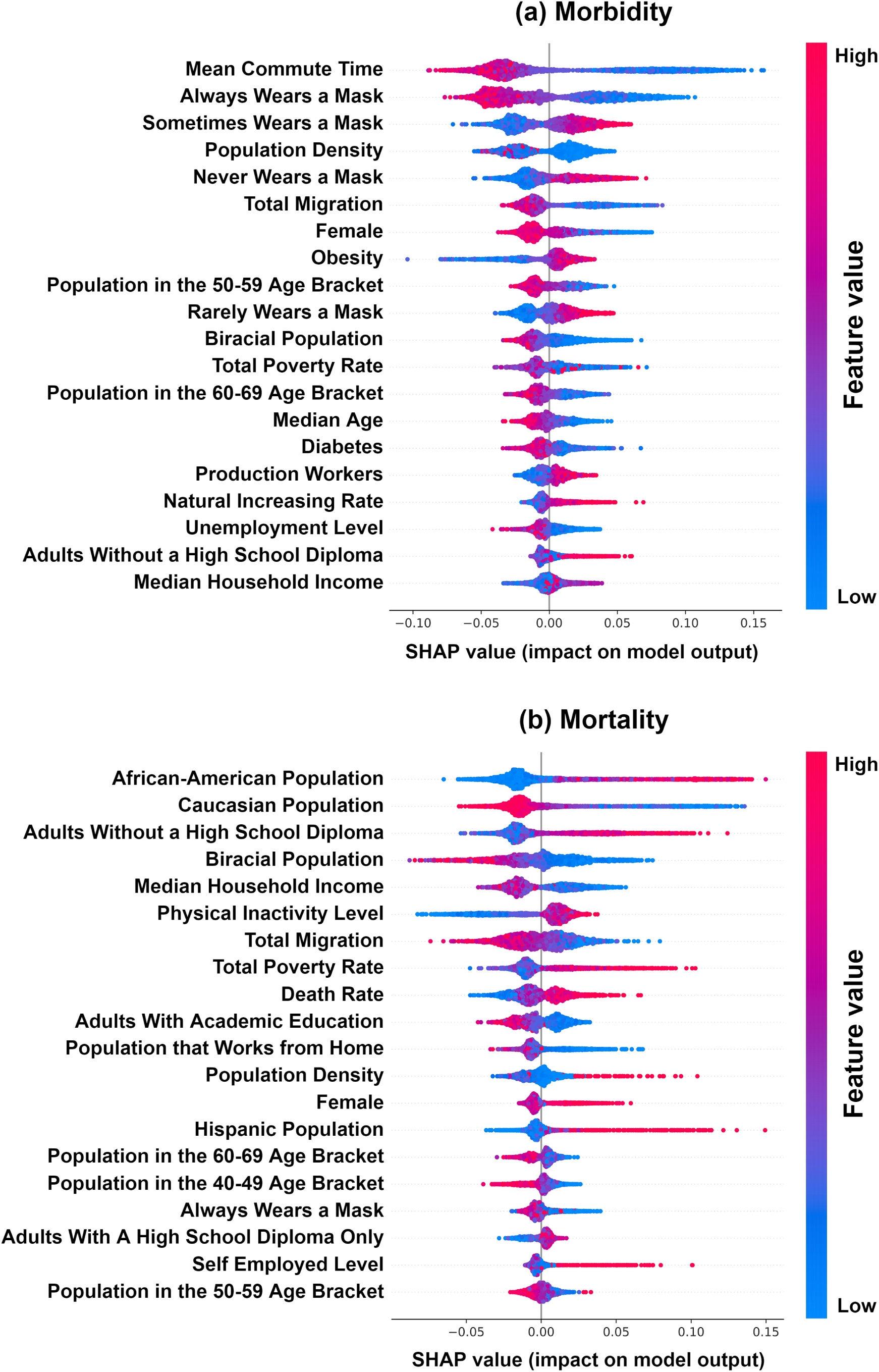
SHAP summary output of the COVID-19 morbidity classification model (a) and mortality classification model (b). Features are sorted by the absolute sum of their SHAP magnitude value, the top features being the most impactful.

The colors correspond to the *value* of each instance of the feature, red being the highest and blue being the lowest. For example, in the case of *mortality* (Fig. 1.b), one can see a county with a very *low* percentage (i.e., colored as deep blue) of Caucasian people, which had produced an impact SHAP value of approximately +0.15. Since +0.15 is a (relatively) large and positive value, that feature contributed to a *high* likelihood for that particular county instance being classified by the model as having a “High Mortality” label.

Three out of the top five highest-impact features within the *morbidity* model (see Fig. 1.a) were related to wearing a mask. “*Always wearing a mask* “correlated *negatively* with morbidity; other mask wearing modes were correlated positively with it.

*Mean Commute Time* had a very high impact on the morbidity model. It was surprisingly *negative*; one would predict that having people travel long distances in public transportation might *increase* COVID-19 cases, as noted in other studies [10].

We shall soon examine such surprising results in more detail.

*Population density*, a feature with a higher value in metropolitan counties and lower for rural ones, also had a surprisingly negative impact on morbidity prediction.

“*Total Migration*” describes how many people entered or left the county in 2019. Like population density, this is a feature associated with larger cities, since they often enjoy positive migration rates (i.e., more people enter the city than leave). Again, the correlation of lower migration rates with higher morbidity seems initially quite surprising.

The percentage of females in the county had a negative impact, in which *higher* values resulted in *lower* morbidity rates, and vice versa.

Obesity had a slightly positive, but mostly a large negative impact, on morbidity.

The production-workers percentage, Natural Increasing [Growth] Rate (the difference between the birth rate and death rate) and percentage of adults without a high school diploma positively affected morbidity.

For the COVID-19 *mortality* model (see SHAP summary output in Fig. 1.b), the most (positively) impacting feature was the African-Americans percentage.

The *education level* of the county’s citizens was an important factor: Next to ethnicity, the most (positively) impacting feature was the rate of adults *without* a high school diploma.

Lower median household incomes and lower total migrations had a negative impact on mortality prediction. As in the case of migration, household income tends to be higher in urban counties [17].

Total poverty rates had a strong positive impact on the mortality model, indicating that counties with a higher percentage of poor citizens suffered more deaths.

The *death rate* feature (the proportional mortality rate that the county had experienced in 2019) had a somewhat high impact.

Working from home, always wearing a mask, and population between 40-69 had a negative impact on the mortality model; population density, female percentage and people who are self employed had a positive impact on it.

Our results suggest a strong influence of ethnicity, socioeconomic state, and self-reported mask-wearing, on county morbidity and especially mortality rates.

However, the other, more surprising results suggest the need for examining the data from an additional aspect: *Time*.

### 3.1 BEHAVIOR OF MORBIDITY AND MORTALITY PREDICTORS OVER TIME

Since the beginning of the pandemic, COVID-19 has been suspected to hit harder the lower-income population, and indeed, most of our results confirm this suspicion.

However, when analyzing, using the same computational methodology (Machine Learning, then calculation of SHAP impact values), the same data types, but at *different time points along 2020*, starting from April 2020, it becomes clear that some features behave similarly even as more data becomes available, while others seem to have an *inverse* impact on morbidity or mortality than the one they started with.

Several socioeconomic features, such as total poverty rates, or percentage of African-Americans in the population seem to have an essentially similar impact on the model’s predictions over time, in this case, a high and positive impact on the morbidity and mortality predictions, respectively.

Other features, however, seem to change over time both their absolute impact, and its *direction* of influence (Fig. 2).

**Fig. 2.**
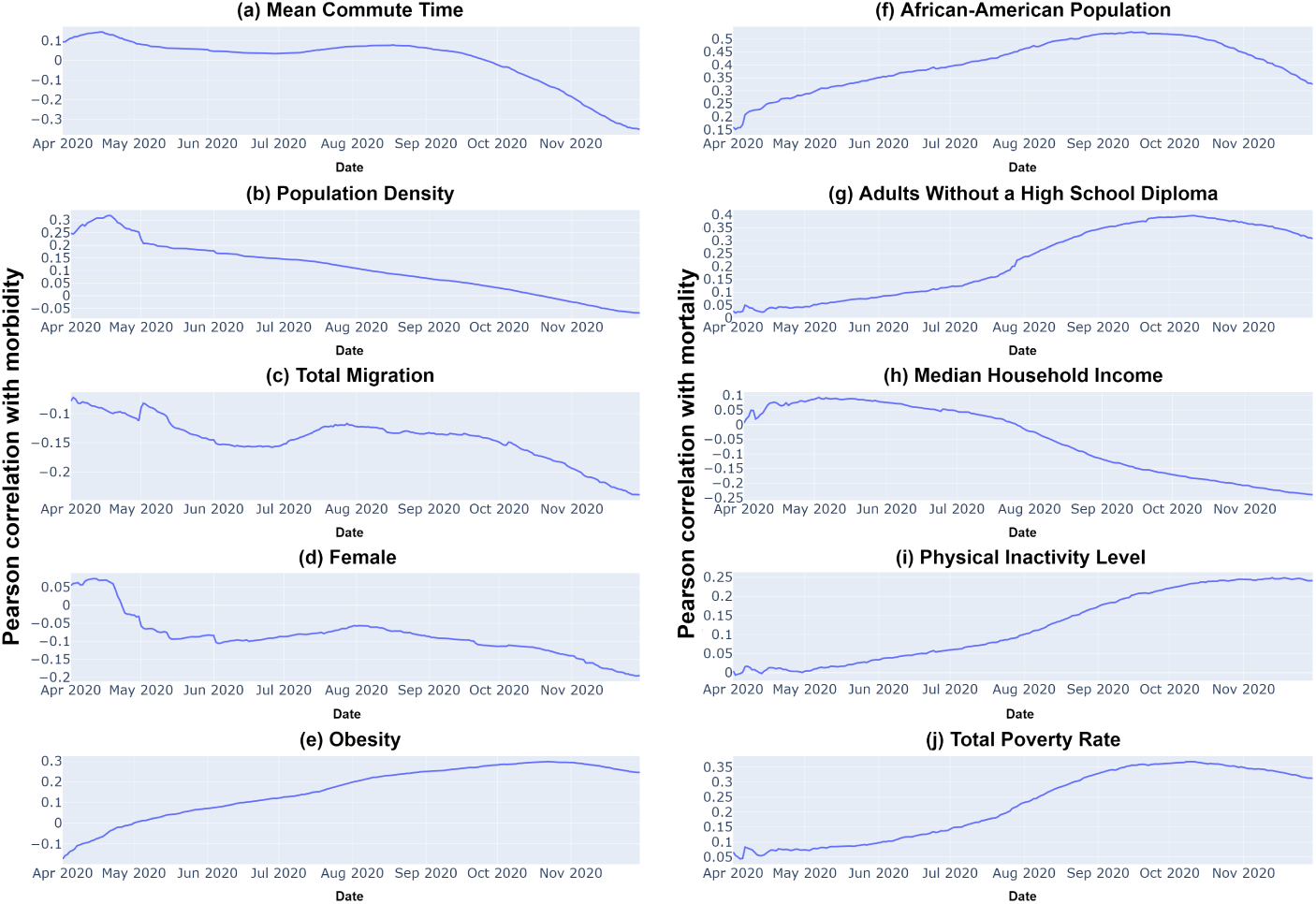
The Pearson correlations between the five most impactful features for each model and the percentage of morbidity (*a* through *e*) and mortality (*f* through *j*). The features are sorted such that the more impactful ones are higher (i.e., *a* and *f* are the most impactful for the morbidity and mortality model, respectively). The correlation is plotted over time, from the 1st of April until the 28th of November.

The most striking “flipping sides” factor is the “*Mean Commute Time*” (Fig. 2.a) - a feature describing the average time it takes for people to arrive at work. In early August 2020, the value of this feature had a low but positive impact on the number of COVID-19 cases. However, when calculated using data collected up to the 28th of November 2020, the correlation is in the opposite direction: now, higher commute times are associated with *lower* morbidity rates), and this feature now provides the highest absolute impact on the morbidity model.

Figure 2.a presents the Pearson correlation, over time, between the “*Mean Commute Time*” feature, and the percentage of COVID-19 cases in the county. We can clearly see the positive correlation with COVID-19 cases *until the beginning of September*. It then decreases quickly and flips to actually become negative, and even considerably so.

We analyzed similarly all features correlated with morbidity and mortality since April 1, 2020 until November 28th, 2020. Several of these are exemplified in Figure 2.

Like the Mean Commute Time, multiple features presented a similar correlation direction (positive or negative) from the 1st of April until approximately *September 1st*. Then, numerous features rapidly decreased their correlation with the morbidity outcome, sometimes even completely flipping the correlation’s direction.

Other features maintained their consistent impact, though it might have changed its magnitude. For example, the correlation between the percentage of African Americans in a county and the mortality reached its peak during September (+0.5) and by November 28th dropped to +0.3 (Fig. 2.f).

Note that the U.S. experienced a surge of COVID-19 cases around mid-October. Combined with the fact that features altered their correlation only slightly earlier, it is likely that a gradual process occurred, which by September had changed the typical profile of the counties that are characterized by relatively high COVID-19 morbidity.

We suspect that this *correlation reversal* is related to how *rural* or *urban* each county is - in the *beginning* of the pandemic, *highly urban* states such as New York and New Jersey bore the brunt of the virus. Later on, less densely populated states such as North Dakota, South Dakota, Iowa and Nebraska had experienced a rise of COVID-19 cases and were found to have the highest numbers, relative to population size.

### Urbanity and Ruralness of Effected Counties Over Time

We can test this theory by referring to a classification scheme used by the U.S. Department of Agriculture Economic Research Service (USDA) named “*Rural-Urban Continuum Codes*” (RUCC) [18]: the USDA assigns each county in the U.S. an RUCC value from 1 to 9, representing how urban or rural it is based on the county’s own population and the population of adjacent counties. An RUCC of one represents the most “urban” counties (metro areas with a population of 1 million or more); an RUCC of nine represents the most “rural” counties (less than 2,500 population, not adjacent to a metro area). Typically, counties with an RUCC of 1 to 3 are considered metropolitan and comprise approximately 37% of the counties in the U.S., while counties with an RUCC of 4 to 9 are considered non-metropolitan and comprise 63% of the counties in the U.S.

The total RUCC distribution can be found in the Supplementary Materials.

Our analysis shows that counties in the top quartile of “mean commute time” values have *lower* RUCC values, with a value of one being predominant.

Other features such as “always wear a mask”, which correlated negatively with morbidity, also characterize mostly metropolitan counties, when focusing on the top quartile of their values.

Figure 3.1 presents from the top down the distribution, at three different time points (April 1st, September 1st, November 28th), of RUCC values for the counties that are at the top quartile of COVID-19 morbidity (Fig. 3.1.a) and deaths, or mortality (Fig. 3.1.b).

**Fig. 3.**
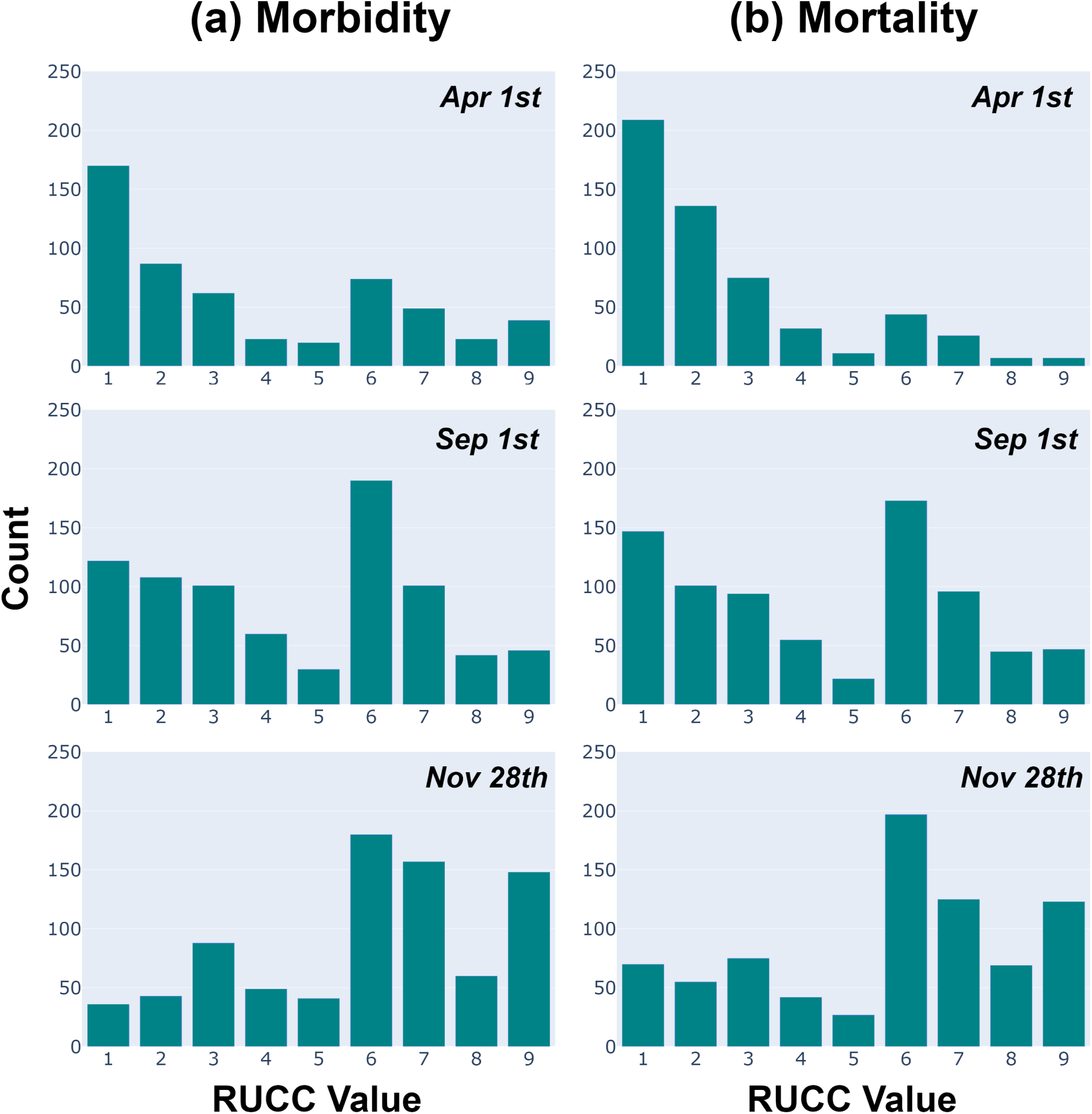
Histograms for the Rural-Urban Continuum Codes (RUCC) distribution of the counties in the top quartile for rate of COVID-19 cases (a) and deaths (b). The vertical axis denotes the number of counties and the horizontal axis the RUCC value. Each row represents a different date. Time progresses from top to bottom.

During early April, both cases and deaths were highly concentrated at the urban counties. However, as time passes, the counties manifesting both the highest morbidity and mortality rates tend to be the more rural ones. *September 1st is a tipping point in which the sum of the urban counties in the top morbidity and mortality quartiles is similar to that of the rural ones*, and by November 28th, the ratio is much higher for rural counties. Clearly, the COVID-19 “wave” spread from the urban counties towards the rural ones.

## 4 DISCUSSION

We can sum up the results of this study, which includes data up to the end of November 2020, with two main conclusions: (1) *The COVID-19 disease is highly correlated with socio-economic status*.

Wealthier counties with fewer minorities, a higher educated population, and lower overall poverty rates had lower morbidity and especially mortality rates. These results are in line with previous research [3,4,2,12,10] using other methods.

(2) *COVID-19 initially affected the most urban areas of the U*.*S. and gradually spread, as a wave, to more rural ones, eventually becoming more prevalent there from September onwards*.

It is also worth noting that obesity is less prevalent in metropolitan counties [19]. Even though the Pearson correlation between obesity and diabetes prevalence is relatively high (0.698), they impacted the model differently. Possibly due to the low variance in diabetes prevalence (5.76 *\** 10^*−*4^) between counties, leading to relatively random results when we compute the SHAP values. In contrast, the variance in obesity prevalence was much higher (2.02 *\** 10^*−*3^).

These results, through a between-counties analysis, confirm in a rather striking fashion previous research that pointed out the discrepancy of mortality rates between different ethnic groups in the U.S. As presented in previous studies [13,15], counties with higher proportions of African American citizens had more COVID-19 cases and deaths. These authors offer multiple explanations and possible reasons for these disparities: occupational type, inadequate access to high-quality medical care, and racial biases in COVID-19 testing and treatment.

Our results suggest that this disparity might well stem from economic reasons.

Several features which are known to increase the mortality from COVID-19 (such as older age, gender, and ICU bed availability) were considered in our study but did *not* appear in the list of highly impacting factors, or appeared with a low weight. In particular, the low impact of the *age* or *gender* features on morbidity and mortality in our results might initially seem surprising, considering the well known association in COVID-19 individual patients between being at an elderly age and suffering the most severe complications; and the higher propensity for death in males.

The likely reason for this lack of association in our current study is a *low variance of these features among counties*, as opposed to their high variance among individuals. The Supplemental Materials demonstrate this observation in more detail.

Other features which were taken into account in the model and in the SHAP computation but which did not appear in the top impacting factors list, include the presidential election results, and the mode of transportation to work.

Note also that COVID-19 morbidity and mortality rates might also be associated with unknown, dynamically changing factors that cannot be easily measured, such as the number of infected but asymptomatic people in each county.

The results in Fig. 2 and Fig. 3.1 present the movement over time of the COVID-19 “wave” across the U.S. This offers an explanation as to why features such as *Mean Commute Time*, which at the *individual* level increases the probability of being infected by COVID-19, actually resulted, by the end of November 2020, in a *negative* correlation with morbidity levels. This is because when viewing such as feature at a county-level, it is actually strongly correlated with the level of *urbanization* (or ruralness) of the county. And *eventually*, the COVID-19 wave hit the rural counties, regarding both morbidity and mortality.

Note that our results do *not* suggest that there is a clear-cut distinction between the urban and rural counties in terms of the *magnitude* of COVID-19 deaths: for example, the total poverty rate is generally higher in *urban* counties [17]; but nevertheless, over the whole period, it had a high positive impact on the mortality model. This seems quite likely, because poverty is a better, more direct predictor for a county’s mortality rate than its level of ruralness or urbanization: Poor people tend to not have access to proper healthcare, whether they live in a sprawling city or in the peripheral countryside.

In addition, the African-American population is distributed rather uniformly across counties with respect to their RUCC [20]; thus, the RUCC value of a county is not enough to predict morbidity or mortality rates; as we had demonstrated, its percentage of African-American population is the strongest predictor of mortality within our model.

Note also that the data regarding COVID-19 cases and deaths used in this study have been collected up to November 28, 2020, while the pandemic is still ongoing; new data have become available, and some correlations might have slightly changed.

In addition, all of the county-level features, aside from the mask-wearing survey and the capacity of ICU beds, were extracted before the beginning of the spread of the COVID-19 and might not have captured some rapid changes that might have occurred up to November 28th 2020.

Finally, all counties were considered equal in the current study, even though their area and population size are different in real-life.

In conclusion, our findings might shed some light on (1) why some counties are prone to a relatively high COVID-19 morbidity or mortality, and (2) how that characterization changed over time in the U.S.

Better understanding of these factors and their temporal dynamics might assist us in better focusing preventive and therapeutic measures, such as vaccination, at the spots in which these factors are most likely to be of benefit.

Our methodology, which is based on explainable machine-learning binary classification models, and exploits game-theoretic principles to calculate the impact of multiple factors, can be easily extended to study the spread of other diseases and other countries.

## Data Availability

See supplementary material for more information.

https://github.com/nytimes/covid-19-data

https://www.cdc.gov

https://www.census.gov/

https://townhall.com/

## Acknowledgments

Nevo Itzhak and Tomer Shahar contributed equally to this work and should be considered as co-first authors.

## Appendix - Supplemental Materials

### DATA SOURCES

The data were extracted from several sources.

Daily reports of cumulative COVID-19 cases and deaths for each county and mask-wearing survey data (250,000 responses between July 2 and July 14, 2020) were taken from The New York Times data site [https://github.com/nytimes/covid-19-data]. Note the COVID-19 data used in this study have been collected up to November 28, 2020. Figure 1 presents the rate of COVID-19 cases [morbidity] (left) and COVID-19 induced deaths [mortality] (right), normalized by the county’s population size, in each U.S. county. Figure 1 was created using the Plotly Express package for Python version 4.4.1 [https://plotly.com/python/plotly-express].

**Figure 1:**
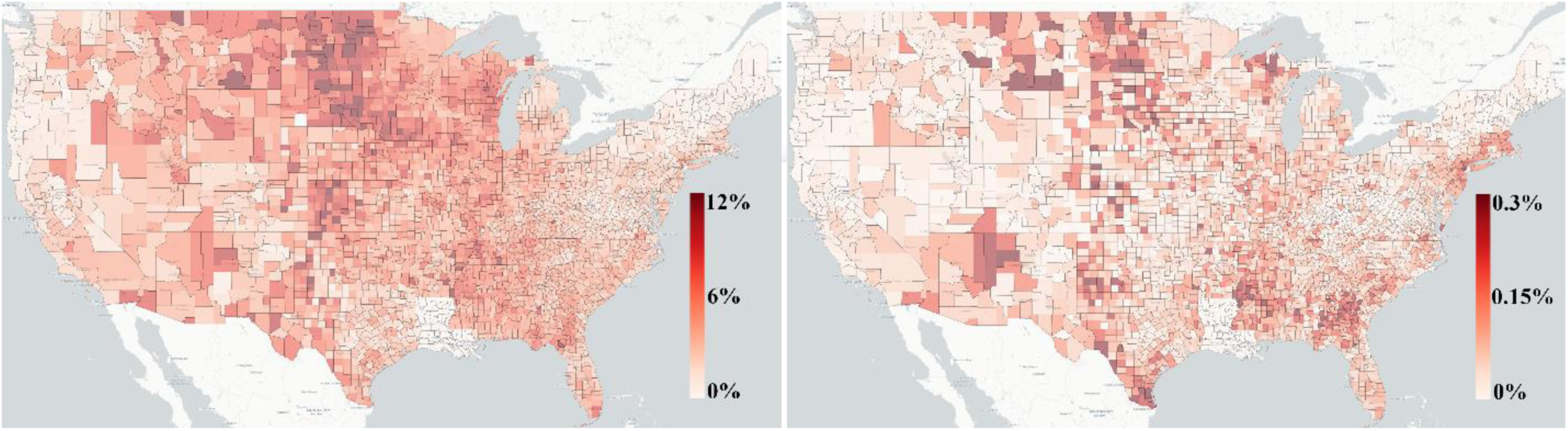
Rate of COVID-19 cases [morbidity] (left) and COVID-19 induced deaths [mortality] (right), normalized by the county’s population size. The color represents the rate of morbidity or mortality, respectively. COVID-19 data have been collected up to November 28, 2020.

Epidemiological factors (diabetes, obesity and inactivity) data (2013) were obtained from the sites of the Centers for Disease Control and Prevention [https://www.cdc.gov].

Ethnicity, socioeconomic, educational attainment, population density and age distribution data (2018) were gathered from the U.S. Census Bureau site [https://www.census.gov].

The socioeconomic data included also the mode of transportation, sector of employment, and ethnicity, amongst other variables. The age dataset contained statistics for each age group featured.

Evaluation of the capacity of ICU, surgical ICU, coronary care unit and burn ICU beds data were taken from the Kaiser Health News site (KHN) [2]. The data were updated up to March 30, 2020 and do not include Veterans Affairs hospitals, since these hospitals do not file cost reports.

County-level presidential election results (in 2016) were taken from the Townhall site [https://townhall.com].

Other than the mask-wearing survey and the capacity of ICU beds, all U.S. county-level features were extracted from repositories created before the beginning of the spread of the COVID-19 pandemic.

The full features list, including their sources and descriptions, can be found in table 1.

**Table 1:** All features used, their source and description.

| Feature Name | Source | Description |
| --- | --- | --- |
| Percent of Adults with Less than a High School Diploma | United States Department of Agriculture (USDA), Economic Research Service | Percent of Adults with Less than a High School Diploma |
| Percent of Adults with a High School Diploma only |  | Percent of Adults with a High School Diploma only |
| Percent of adults that completed some College or Associates Degree |  | Percent of adults that completed some College or Associates Degree |
| Percent of Adults with a Bachelors Degree or Higher |  | Percent of Adults with a Bachelors Degree or Higher |
| Total Poverty Rate |  | Percent of population that are impoverished |
| Death Rate |  | Percent of population that died in 2019 |
| Natural Increasing Rate |  | Increase in percentage of population size in 2019 |
| Total Migration |  | Number of people immigrating/emigrating to the county as a percentage of total population |
| Unemployment Level |  | Percentage of unemployed people |
| Median Household Income |  | Median income of county |
| Diabetes | Living Atlas of the World - Diabetes, Obesity, and Inactivity by US County | Percentage of people with diabetes in the county |
| Physical Inactivity Level |  | Percentage of people that do not perform physical activity |
| Obesity |  | Percentage of obese people in the county |
| Female | United States Census Bureau | Percentage of females in the county |
| African-American Population |  | Percentage of African-Americans in the county |
| Caucasian Population |  | Percentage of Caucasians in the county |
| Asian Population |  | Percentage of Asians in the county |
| Hispanic Population |  | Percentage of Hispanics in the county |
| American Indian Population |  | Percentage of Indians in the county |
| Biracial Population |  | Percentage of people with mixed heritage in the county |
| ICU Beds per person | Kaiser Health News (KHN) | Number of beds in ICU wards per person |
| Percentage of Democratic Voters | https://townhall.com/ | Percent of people that voted for the Democrats in the 2016 elections |
| Voting Difference |  | Absolute difference between percent of Democratic and Republican voters for the 2016 elections |
| Voting Turnout |  | Percent of eligible voters in the 2016 elections (out of the whole population) |
| Professional Workers | United States Census Bureau | Percent of population that provide professional services |
| Service Workers |  | Percent of population that provide government services |
| Office Workers |  | Percent of population that work in offices in the private sector |
| Construction Workers |  | Percent of population that work in construction work |
| Production Workers |  | Percent of population that work in production facilities (e.g., factories) |
| Drive to Work |  | Percent of population that commutes by driving |
| Carpools to Work |  | Percent of population that commutes by carpooling |
| Public Transportation to Work |  | Percent of population that commutes with public transportation |
| Walk to Work |  | Percent of population that commutes by walking |
| Other Transportation to Work |  | Percent of population that commutes by other means |
| Population that Works from Home |  | Percent of population that works from home |
| Mean Commute Time |  | Average amount of time it takes people to get to work (in hours) |
| Private Work Sector |  | Percent of people that work in the private sector |
| Public Work Sector |  | Percent of people that work in the public sector |
| Self Employed |  | Percent of people that are self-employed |
| Family Work Sector |  | Percent of people that work in a family business |
| Never Wears a Mask | New York Times<br>Github Repository | Percent of people that never wear a mask |
| Rarely Wears a Mask |  | Percent of people that rarely wear a mask |
| Sometimes Wears a Mask |  | Percent of people that sometimes wear a mask |
| Frequently Wears a Mask |  | Percent of people that frequently wear a mask |
| Always Wears a Mask |  | Percent of people that always wear a mask |
| Population Density | United States<br>Census Bureau | Number of people per square mile in the county |
| Median Age | United States<br>Department of<br>Agriculture (USDA),<br>Economic Research<br>Service | Median age of people in the county |
| Population in the 20-29 age bracket |  | Percent of population between the ages of 20-29 |
| Population in the 30-39 age bracket |  | Percent of population between the ages of 30-39 |
| Population in the 40-49 age bracket |  | Percent of population between the ages of 40-49 |
| Population in the 50-59 age bracket |  | Percent of population between the ages of 50-59 |
| Population in the 60-69 age bracket |  | Percent of population between the ages of 60-69 |
| Population in the 70+ age bracket |  | Percent of population over the age of 70 |

### DATA PRE-PROCESSING METHODS

We merged the described datasets, using *Federal Information Processing Standard Publication* (FIPS) counties codes, and removed features that were highly correlated, either positively or negatively (absolute Pearson correlation coefficient greater than 0.9), using only one feature from each set of correlated features. There were no missing values in the final merged data set.

The features were normalized into a range of zero and one, aside from natural increasing rate, death rate, and total migration. These three features have negative values. Thus, we normalized these features into a range of minus one and one. Table 2 presents the features and their statistics: mean, standard deviation (STD), minimum (Min), the first quartile (Q1), median, the third quartile (Q3), and the maximum (Max). For example, to normalize the *Mean Commute Time* feature to have a value between zero and one, we divided it by 24 hours. This ensures the feature value will be in that range. Similarly, to normalize the *Mean Age* feature into to have a value between zero and one, we divided it by 120 years, since 120 is much greater than even the maximal value to be found for that feature.

**Table 2:** Descriptive statistics for all features: mean, standard deviation (STD), minimum (Min), the first quartile (Q1), median, the third quartile (Q3), and the maximum (Max).

| Feature Name | Mean | STD | Min | Q1 | Median | Q3 | Max |
| --- | --- | --- | --- | --- | --- | --- | --- |
| Percent of Adults with Less than a High School Diploma | 0.134 | 0.063 | 0.012 | 0.088 | 0.121 | 0.172 | 0.663 |
| Percent of Adults with a High School Diploma only | 0.344 | 0.071 | 0.081 | 0.299 | 0.347 | 0.394 | 0.556 |
| Percent of adults that completed some College or Associates Degree | 0.308 | 0.052 | 0.058 | 0.273 | 0.307 | 0.342 | 0.573 |
| Percent of Adults with a bachelor's degree or Higher | 0.214 | 0.092 | 0 | 0.15 | 0.191 | 0.254 | 0.746 |
| Total Poverty Rate | 0.151 | 0.061 | 0.026 | 0.108 | 0.141 | 0.182 | 0.484 |
| Death Rate | -0.055 | 0.238 | -1 | -0.201 | -0.048 | 0.107 | 1 |
| Natural Increasing Rate | -0.261 | 0.204 | -1 | -0.393 | -0.273 | -0.147 | 1 |
| Total Migration | 0.137 | 0.085 | -1 | 0.098 | 0.134 | 0.176 | 1 |
| Unemployment Level | 0.176 | 0.076 | 0 | 0.124 | 0.161 | 0.21 | 0.946 |
| Median Household Income | 0.237 | 0.119 | 0 | 0.159 | 0.219 | 0.289 | 1 |
| Diabetes | 0.113 | 0.025 | 0.033 | 0.096 | 0.111 | 0.129 | 0.235 |
| Physical Inactivity Level | 0.26 | 0.052 | 0.081 | 0.227 | 0.259 | 0.295 | 0.414 |
| Obesity | 0.311 | 0.045 | 0.118 | 0.284 | 0.312 | 0.339 | 0.476 |
| Female | 0.499 | 0.022 | 0.268 | 0.494 | 0.503 | 0.51 | 0.569 |
| African-American Population | 0.091 | 0.143 | 0 | 0.009 | 0.025 | 0.104 | 0.861 |
| Caucasian Population | 0.851 | 0.157 | 0.087 | 0.804 | 0.916 | 0.956 | 0.99 |
| Asian Population | 0.015 | 0.027 | 0 | 0.005 | 0.007 | 0.014 | 0.43 |
| Hispanic Population | 0.096 | 0.139 | 0.006 | 0.024 | 0.043 | 0.1 | 0.964 |
| American Indian Population | 0.021 | 0.067 | 0 | 0.004 | 0.006 | 0.013 | 0.857 |
| Biracial Population | 0.021 | 0.014 | 0 | 0.013 | 0.018 | 0.024 | 0.303 |
| ICU Beds per person | $10^{-4}$ | $5 \cdot 10^{-3}$ | 0 | $<10^{-6}$ | $<10^{-6}$ | $<10^{-6}$ | 0.028 |
| Percentage of Democratic Voters | 0.313 | 0.149 | 0.031 | 0.204 | 0.283 | 0.394 | 0.893 |
| Voting Difference | 0.392 | 0.207 | 0 | 0.224 | 0.405 | 0.554 | 0.916 |
| Voting Turnout | 0.75 | 0.052 | 0.457 | 0.732 | 0.76 | 0.782 | 0.907 |
| Professional Workers | 0.314 | 0.064 | 0.114 | 0.273 | 0.305 | 0.349 | 0.69 |
| Service Workers | 0.181 | 0.036 | 0 | 0.157 | 0.177 | 0.2 | 0.464 |
| Office Workers | 0.218 | 0.03 | 0.048 | 0.199 | 0.22 | 0.238 | 0.372 |
| Construction Workers | 0.127 | 0.041 | 0.033 | 0.099 | 0.122 | 0.149 | 0.364 |
| Production Workers | 0.16 | 0.058 | 0 | 0.118 | 0.156 | 0.197 | 0.487 |
| Drive to Work | 0.799 | 0.065 | 0.185 | 0.774 | 0.81 | 0.84 | 0.972 |
| Carpools to Work | 0.099 | 0.029 | 0 | 0.081 | 0.095 | 0.113 | 0.293 |
| Public Transportation to Work | 0.008 | 0.021 | 0 | 0.001 | 0.003 | 0.007 | 0.424 |
| Walk to Work | 0.03 | 0.031 | 0 | 0.014 | 0.022 | 0.038 | 0.497 |
| Other Transportation to Work | 0.015 | 0.012 | 0 | 0.008 | 0.013 | 0.019 | 0.211 |
| Population that Works from Home | 0.048 | 0.031 | 0 | 0.029 | 0.041 | 0.058 | 0.33 |
| Mean Commute Time | 0.016 | 0.004 | 0.004 | 0.014 | 0.016 | 0.019 | 0.031 |
| Private Work Sector | 0.752 | 0.073 | 0.321 | 0.718 | 0.763 | 0.803 | 0.888 |
| Public Work Sector | 0.167 | 0.059 | 0.044 | 0.126 | 0.156 | 0.193 | 0.623 |
| Self Employed | 0.078 | 0.039 | 0 | 0.053 | 0.068 | 0.092 | 0.38 |
| Family Work Sector | 0.003 | 0.005 | 0 | 0.001 | 0.002 | 0.003 | 0.08 |
| Never Wears a Mask | 0.081 | 0.059 | 0 | 0.034 | 0.069 | 0.115 | 0.432 |
| Rarely Wears a Mask | 0.084 | 0.056 | 0 | 0.041 | 0.074 | 0.116 | 0.384 |
| Sometimes Wears a Mask | 0.122 | 0.058 | 0.003 | 0.08 | 0.116 | 0.157 | 0.422 |
| Frequently Wears a Mask | 0.207 | 0.062 | 0.029 | 0.164 | 0.204 | 0.247 | 0.549 |
| Always Wears a Mask | 0.506 | 0.152 | 0.115 | 0.392 | 0.496 | 0.611 | 0.889 |
| Population Density | 0.002 | 0.007 | 0 | 0 | 0 | 0.001 | 0.172 |
| Median Age | 0.349 | 0.044 | 0.194 | 0.322 | 0.348 | 0.373 | 0.568 |
| Population in the 20-29 age bracket | 0.121 | 0.03 | 0.05 | 0.104 | 0.117 | 0.13 | 0.373 |
| Population in the 30-39 age bracket | 0.118 | 0.017 | 0.058 | 0.107 | 0.116 | 0.126 | 0.211 |
| Population in the 40-49 age bracket | 0.115 | 0.013 | 0.059 | 0.107 | 0.115 | 0.123 | 0.179 |
| Population in the 50-59 age bracket | 0.133 | 0.014 | 0.047 | 0.125 | 0.134 | 0.142 | 0.189 |
| Population in the 60-69 age bracket | 0.134 | 0.025 | 0.035 | 0.118 | 0.132 | 0.147 | 0.261 |
| Population in the 70+ age bracket | 0.136 | 0.035 | 0.036 | 0.113 | 0.133 | 0.155 | 0.439 |

Please note the statistics in table 2 are for the 3,071 counties that were used in the main paper and not all of the counties in the U.S.

### COMPUTATIONAL METHODS

Figure 2 displays a histogram of the COVID-19 mortality rate and COVID-19 morbidity rate in the U.S. The top quartile thresholds were used to determine the class for each county for either COVID-19 morbidity or mortality.

**Figure 2:**
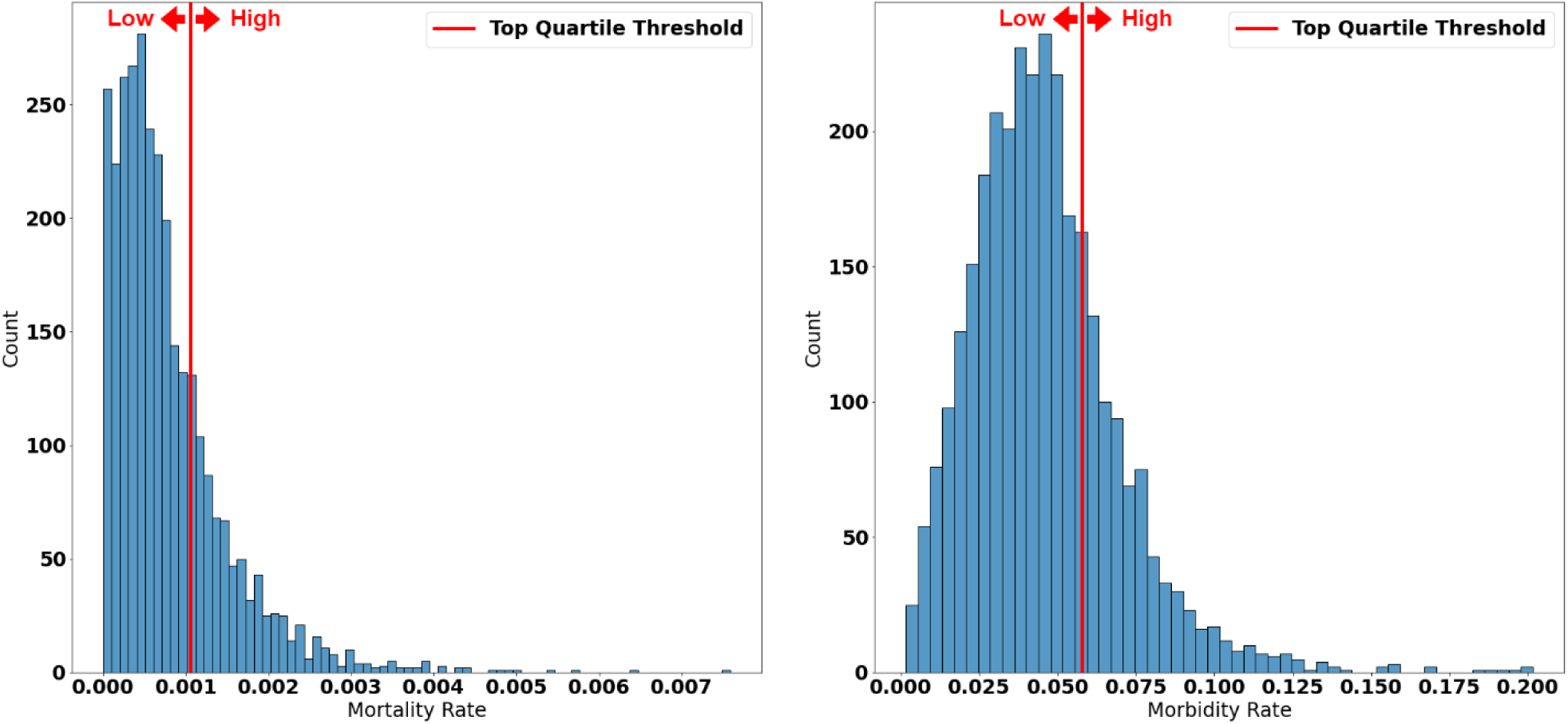
Histogram of the COVID-19 mortality rate (left) and COVID-19 morbidity rate (right). The red vertical line represents the top quartile thresholds.

There was no direct correlation between the morbidity and mortality rates in each county.

For both models, we used a Random Forest algorithm to induce the classifier without bootstrapping. The parameters of each model were selected after testing each combination’s performance using a grid search approach. The grid search approach used ten cross-validations to select the best parameters.

The minimum number of samples required to split an *internal* node was 2, with a maximal depth of 15; and the random-generator seed was 2020.

For the mortality model, the minimum number of samples required to be at a *leaf* node was 5 and the number of trees in the forest was 200. For the morbidity model, the minimum number of samples required to be at a *leaf* node was 4 and the number of trees in the forest was 600. For the parameters we did not specify, we used the default. Random Forest classifier and Grid search approach was implemented with Python 3.7 Scikit-Learn version 0.22.2 [https://scikit-learn.org/].

Once the model converged, and we confirmed that it achieved a reasonable accuracy, the SHAP approach was used for analyzing the impact of each feature on the testing set. It is necessary to verify that the models are accurate enough. Otherwise, the feature importance is not significant enough to draw conclusions from. We achieved fairly high AUC and AUPRC scores. Thus, both models are sufficiently accurate for the impact weights to be meaningful. To coverage all U.S. counties, we merge the SHAP values from ten different iteration into a one figure.

This algorithm is a way to reverse-engineer the output of any predictive algorithm. SHAP values are used when there is a complex model and we want to understand what decisions the model is making. In general, the SHAP values show how much a given feature changed our prediction and in which direction. SHAP was implemented using the SHAP package for Python version 0.35.0 [https://github.com/slundberg/shap].

### EVALUATION METRICS

This section describes the evaluation metrics we used, using as an example the distinction method we chose for the mortality rate; the morbidity rate was divided similarly.

The *True Positive Rate* (TPR) (also referred to as *sensitivity* or *recall*) is the proportion of counties who were classified by the model as being “High Mortality” (true positives) out of counties who were labeled as being in the “High Mortality” subset. The TPR represents the model’s ability to find all “High Mortality” counties.

The *False Positive Rate* (FPR) (also referred to as the *specificity*) is the proportion of counties wrongly categorized as “High Mortality” (false positives) out of the total number of labeled as “Low Mortality”. The FPR represents the probability of a false alarm - i.e., incorrectly predicting a “High Mortality” label.

The *Precision* (also referred to as the *Positive Predictive Value* (PPV) is the proportion of counties who were (correctly) labeled as belonging to the “High Mortality” subgroup, out of those who were classified as belonging to the “High Mortality” subgroup. The precision is the model’s accuracy when predicting the “High Mortality” label for a given county.

The *Receiver Operating Characteristic* (ROC) graph displays on the vertical axis the TPR, and on the horizontal axis the FPR, for all of the binary classification thresholds.

The *Precision-Recall* graph displays on the vertical axis the recall (i.e., the TPR) and on the horizontal axis the precision (PPV) for different probability thresholds.

To quantify the classification performance of both the morbidity and mortality models, we used two evaluation metrics: (1) the *Area Under the ROC curve* (AUROC), and (2) the A*rea Under the Precision-Recall Curve* (AUPRC). Higher AUROC and AUPRC values imply that the model is more accurate. Interested readers might wish to read more about these metrics and the relationship between them elsewhere [1].

## RESULTS

Figures 3 and 4 present the twenty most impactful features (vertical axis) on the COVID-19 morbidity and mortality model, versus the rate of COVID-19 cases and deaths (horizontal axis), respectively.

**Figure 2:**
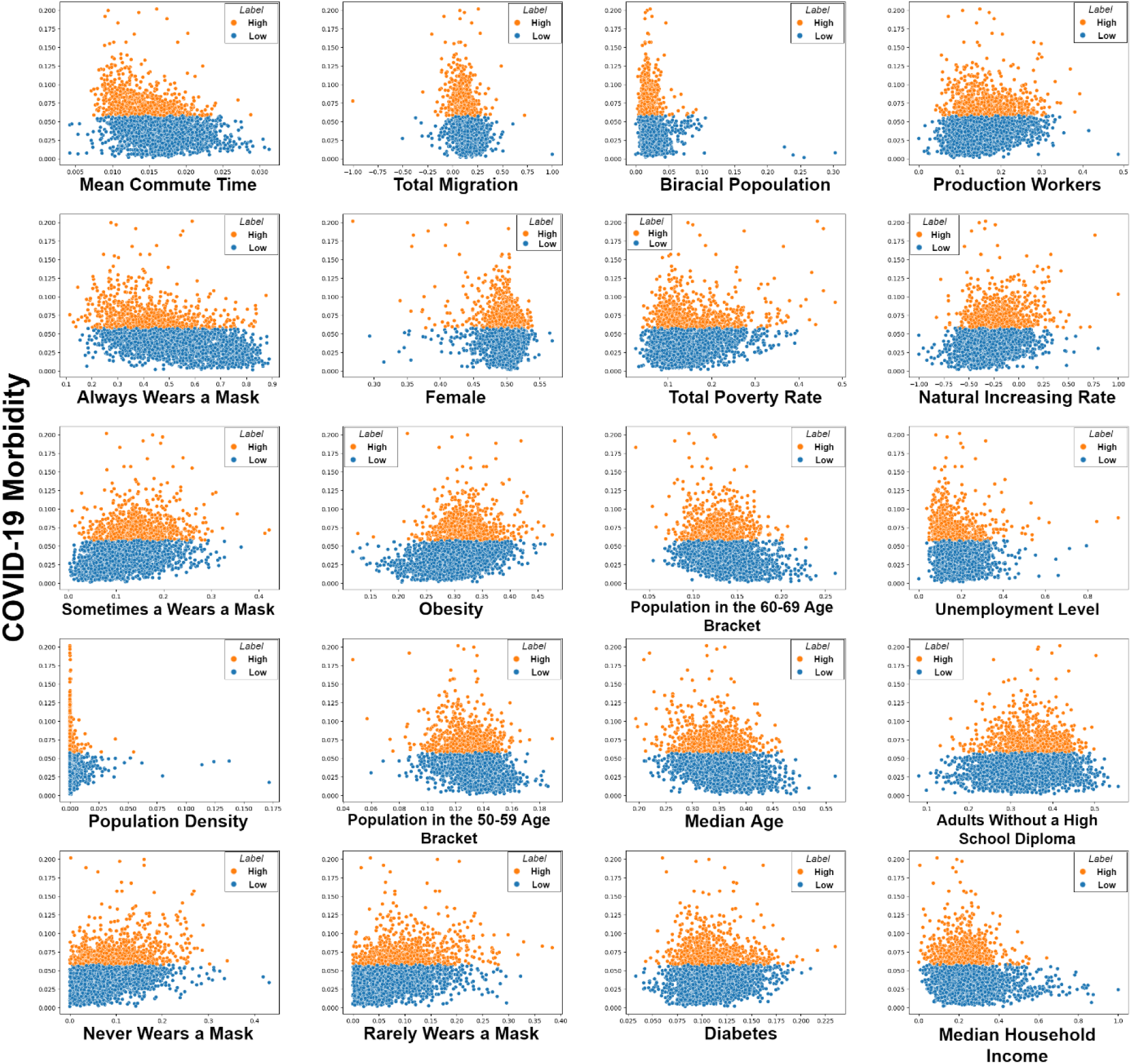
The twenty most impactful features on the COVID-19 morbidity model. Each graph represents the variable values (vertical axis) versus the COVID morbidity rate (horizontal axis) in each county. Orange points represent counties with high COVID-19 morbidity and blue points represent counties with low COVID-19 morbidity counties.

Figure 3 present a variable (vertical axises) versus the rate COVID-19 cases (horizontal axises) in U.S. Orange points represent counties with high COVID19 morbidity, and blue points represent counties with low COVID-19 morbidity counties. The variables order follows the order presented in the results figure in the paper. For example, the upper left variable (i.e., *Mean Commute Time*) is the most impactful factor in the morbidity model followed by the feature *Always Wears a Mask*. The feature *Total Migration* is the sixth most impactful feature and *Median Age* is the fourteenth most impactful feature. As can be seen from the upper left plot, *Mean Commute Time* versus COVID-19 cases, there is a slight negative connection between the *Mean Commute Time* and COVID-19 cases. This negative correlation fits with the results presented in the paper, namely that high values of *Mean Commute Time* led to lower COVID-19 morbidity rates in a county.

**Figure 3:**
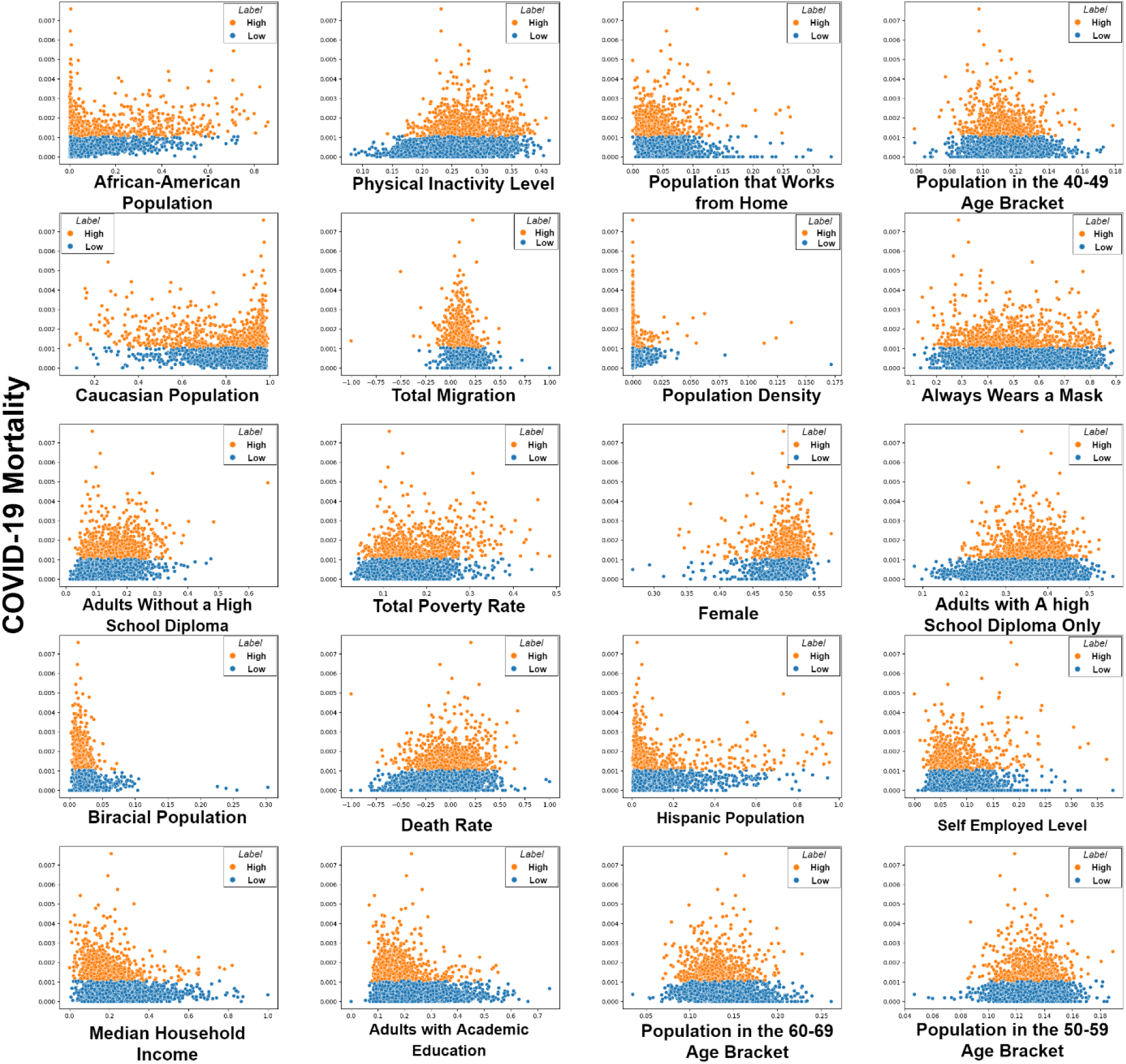
The twenty most impactful features on the COVID-19 mortality model. Each graph represents the variable values (vertical axis) versus the COVID mortality rate (horizontal axis) in each county. Orange points represent counties with high COVID-19 mortality and blue points represent counties with low COVID-19 mortality counties.

The same semantics are present for figure 4 with COVID-19 mortality. For example, the upper left variable (i.e., *African-American Population*) is the most impactful factor in the mortality model, followed by the feature *Caucasian Population*. The feature *Physical Inactivity Level* is the sixth most impactful feature, and *Hispanic Population* is the fourteen-the most impactful feature. As can be seen from the upper left plot, *African-American Population* feature’s plot versus the COVID-19 deaths, there is a slightly positive connection between the *African-American Population* feature and the COVID-19 deaths. This positive correlation fits in with the results presented in the paper, namely that high values of the *African-American Population* feature led to higher COVID-19 morality rates in a county.

**Figure 4:**
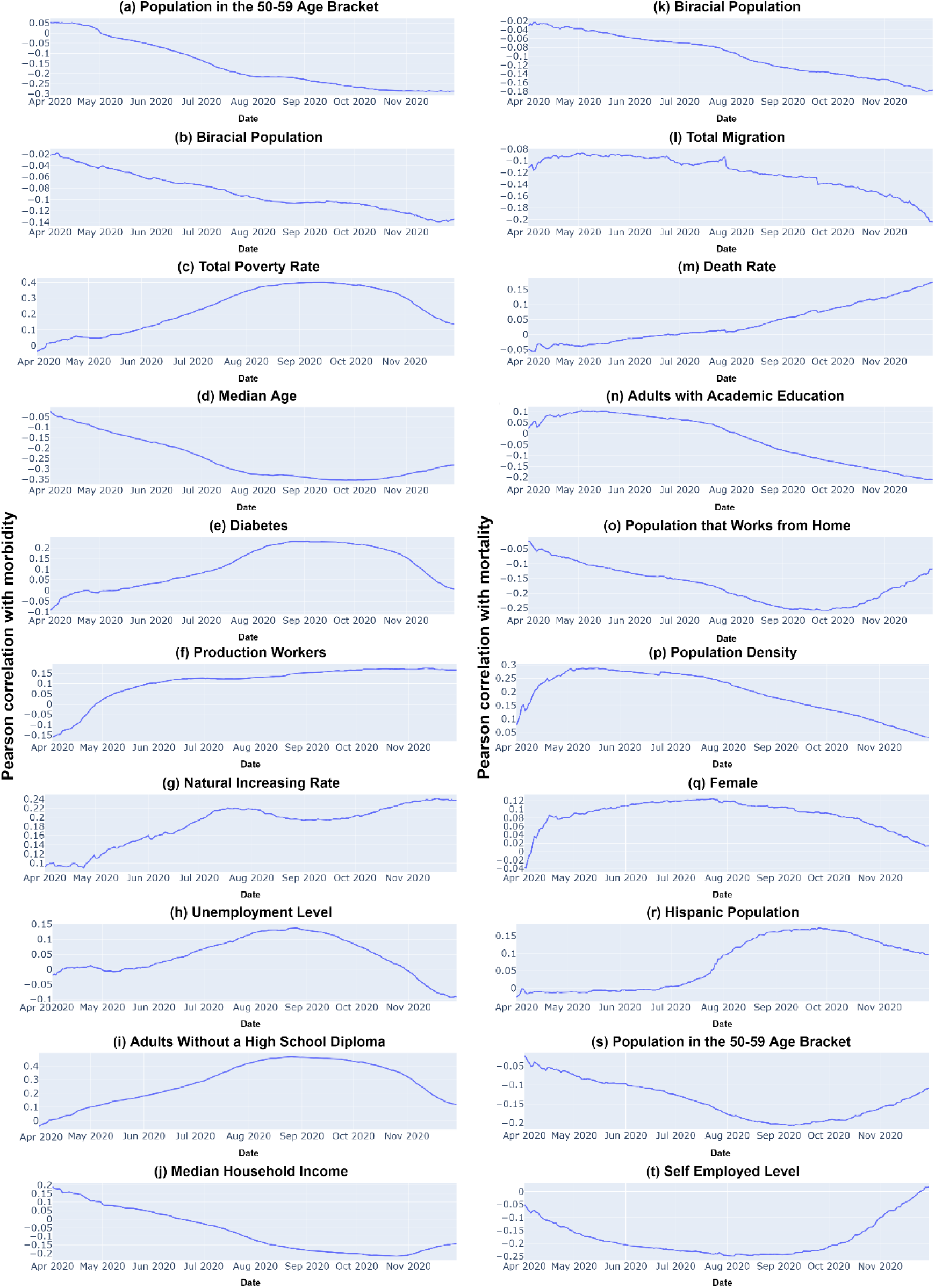
The Pearson correlations between an additional nine features (outside of the ones shown in the main paper) for each model and the percentage of morbidity (a through j) and mortality (k through t). The features are sorted such that the more impactful ones are higher (i.e., a, j have more impact on the morbidity and mortality model than b, l, respectively). The correlation is plotted over time, from the 1st of April until the 28th of November.

### BEHAVIOR OF MORBIDITY AND MORTALITY PREDICTORS OVER TIME

Some features seem to change over time, both their absolute impact and their *direction* of influence. We analyzed all features correlated with morbidity and mortality from April 1, 2020, until November 28, 2020. Figure 5 contains eighteen more Pearson correlation plots, over time, between variables for each model. This figure is provided in addition to the ten plots provided in the paper.

For example, in figure 5.c, in early August 2020, the value of *Total Poverty Rate* feature had a high positive (+0.35) impact on the number of COVID-19 cases. However, when calculated using data collected up to November 28, 2020, the correlation is almost twice less (+0.15).

**Figure 5:**
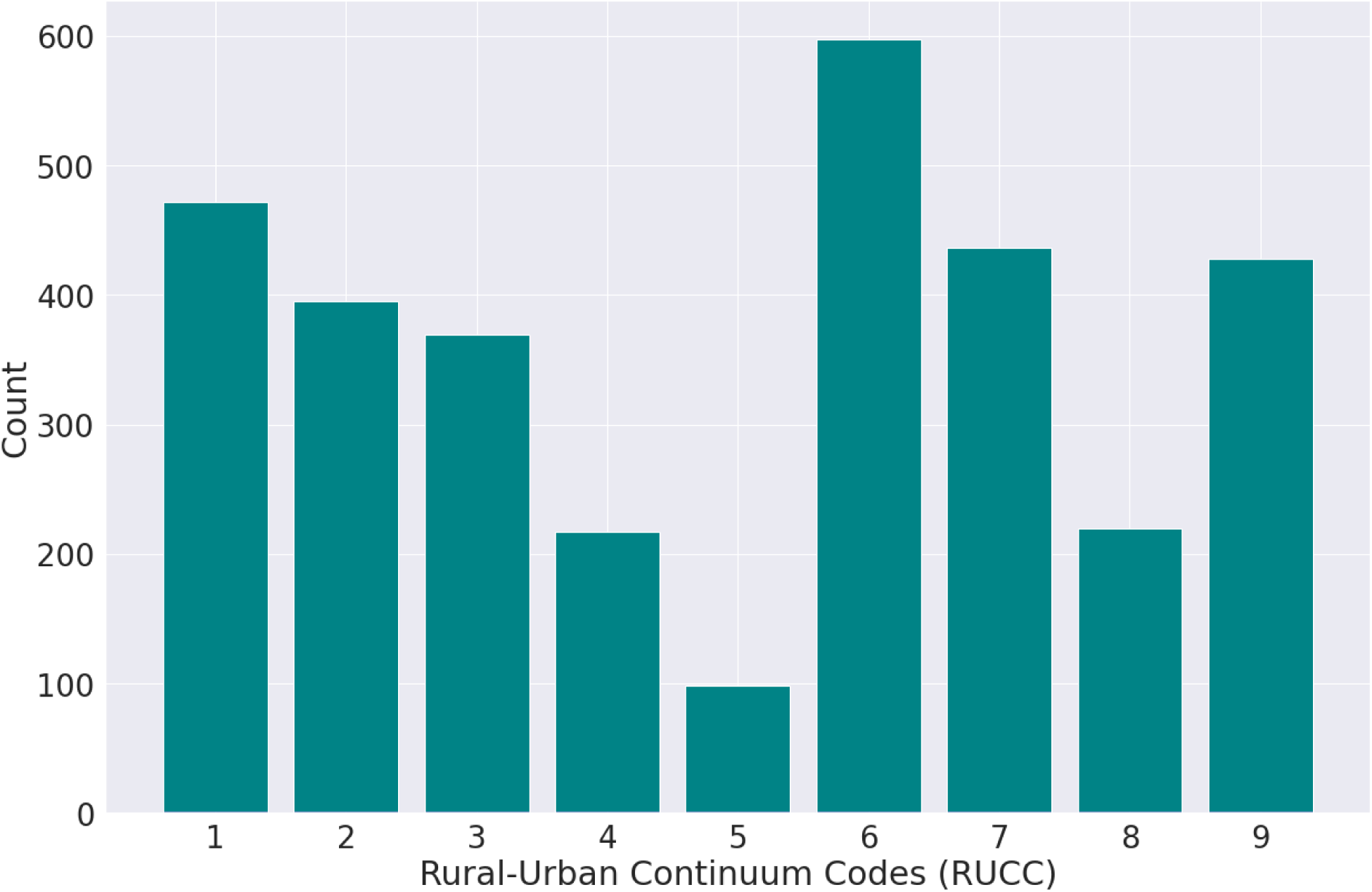
Distribution of Rural-Urban Continuum Codes (RUCC) values in the U.S. The horizontal axis denotes the number of counties, the vertical axis the RUCC value.

### Urbanity and Ruralness of Effected Counties Over Time

The total “*Rural-Urban Continuum Codes*” (RUCC) distribution can be seen in figure 6. The USDA assigns each county in the U.S. an RUCC value from 1 to 9, representing how urban or rural it is based on the county’s own population and the population of adjacent counties.

An RUCC of one represents the most “urban” counties (metro areas with a population of 1 million or more); an RUCC of nine represents the most “rural” counties (less than 2,500 population, not adjacent to a metro area). Typically, counties with an RUCC of 1 to 3 are considered metropolitan and comprise approximately 37% of the counties in the U.S., while counties with an RUCC of 4 to 9 are considered non-metropolitan and comprise 63% of the counties in the U.S.

Figure 7 presents the distribution of RUCC values between counties in the top quartile for the 32 different variables value. The horizontal axis denotes the number of counties, the vertical axis the RUCC value. For example, the upper left plot in figure 7 presents the histogram of the top quartile counties of *Mean Commute Time* values. As can be seen from this histogram, the top quartile counties of *Mean Commute Time* values are more metropolitan counties, which is reasonable. Similar behavior can be found in other features, such as *Always Wears a Mask, Population Density, Adults with Academic Education, Median Household Income*.

**Figure 6:**
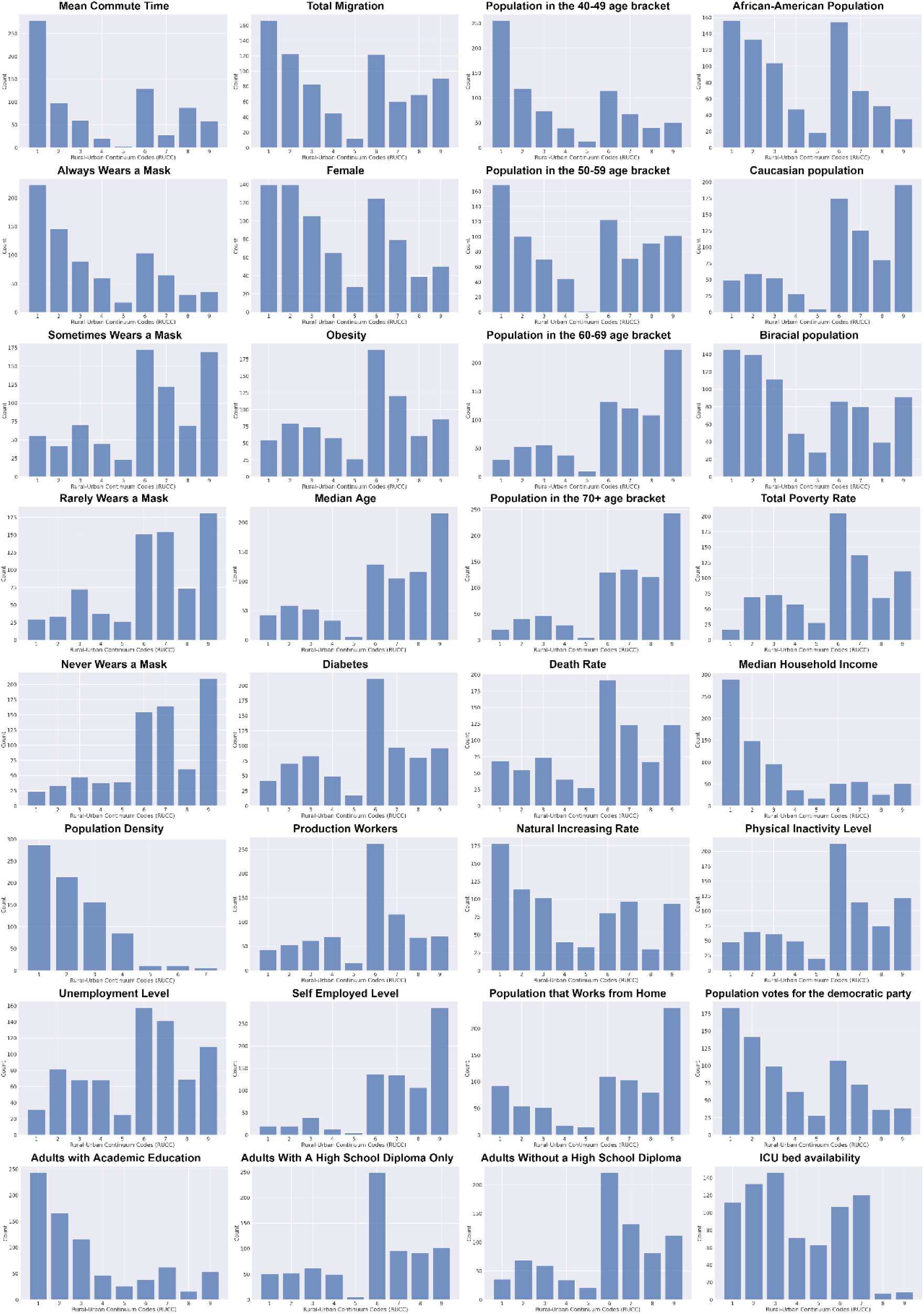
Distribution of Rural-Urban Continuum Codes (RUCC) values between counties that are in the top quartile for the different variables value. The horizontal axis denotes the number of counties, the vertical axis the RUCC value.

### Effect of the Same factors at an Individual Level versus at a Population Level

Several features which are known to increase the mortality from COVID-19 (such as older age, gender, and ICU bed availability) were considered in our study but did *not* appear in the list of highly impacting factors, or appeared with a low impact weight. In particular, the low impact of the *age* features on morbidity and mortality in our results might initially seem surprising, considering the well known association in COVID-19 individual patients between being at an elderly age and suffering the most severe complications; and the higher propensity for death in males.

The likely reason for this lack of association in our current study is a *low variance of these features among counties*, as opposed to their high variance among individuals.

Figure 8 presents box plots for the various age brackets used in our models. As can be seen, there is little variance between almost all of these features. There are a few outliers, such as population in the 20-29 and 70+ bracket. Notice that the quartiles are spread out rather evenly.

**Figure 7:**
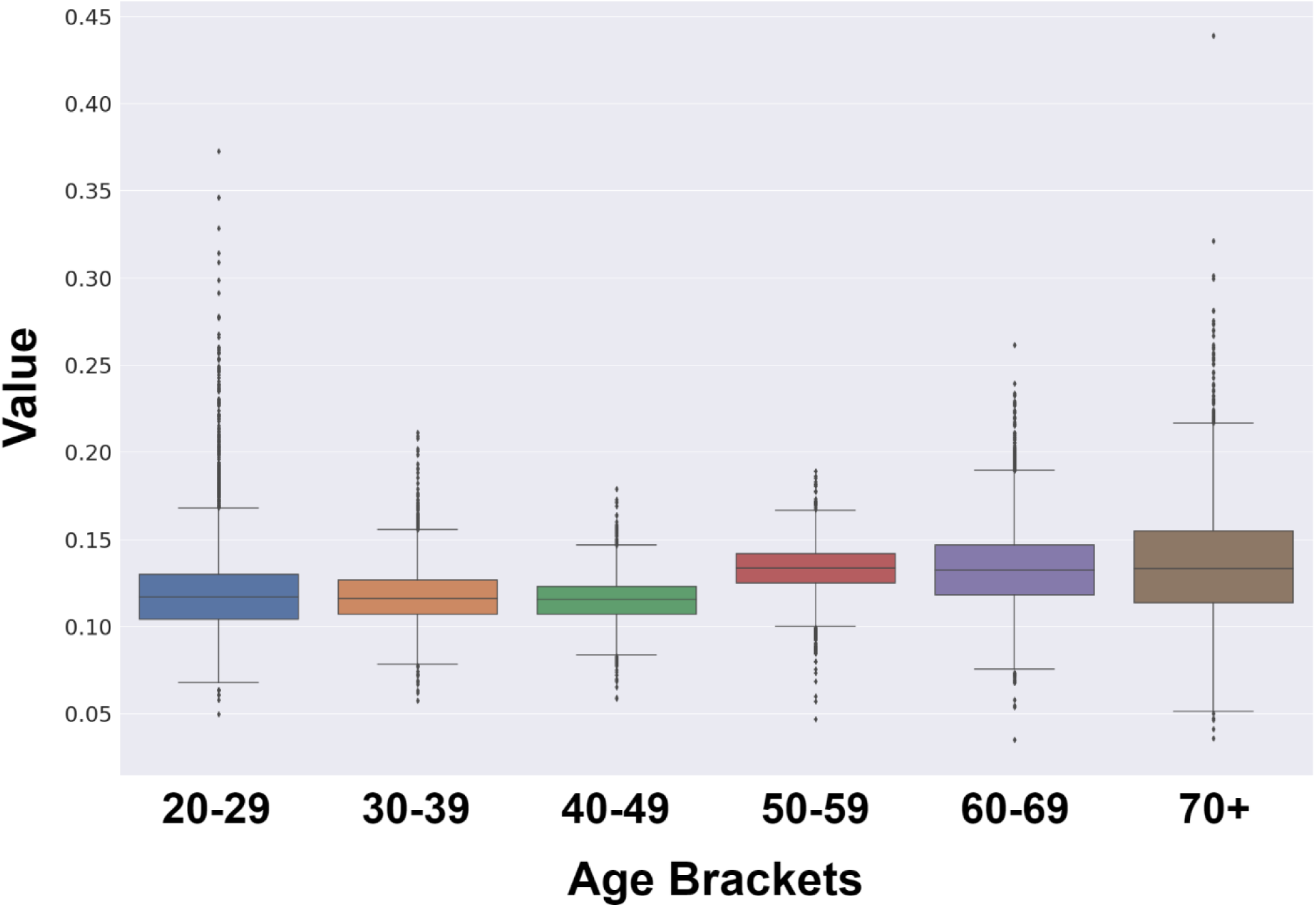
Box plots for African-American population percentage and for Caucasians’ percentage across counties.

In contrast, figure 9 presents box plots for the percentage of Caucasian and African-American populations across counties. We can see a very high variance inside each feature, and a large difference between these features themselves. This is likely to be the reason that age did not emerge in our methodology as one of the top predictive features for morbidity or mortality: for an *individual*, age is a highly significant predictor of the outcome. But when predicting the effect of COVID-19 for a whole *county*, the age distribution withinn the county is not a very useful predictor, because the distribution of age values is quite similar between different counties.

**Figure 8:**
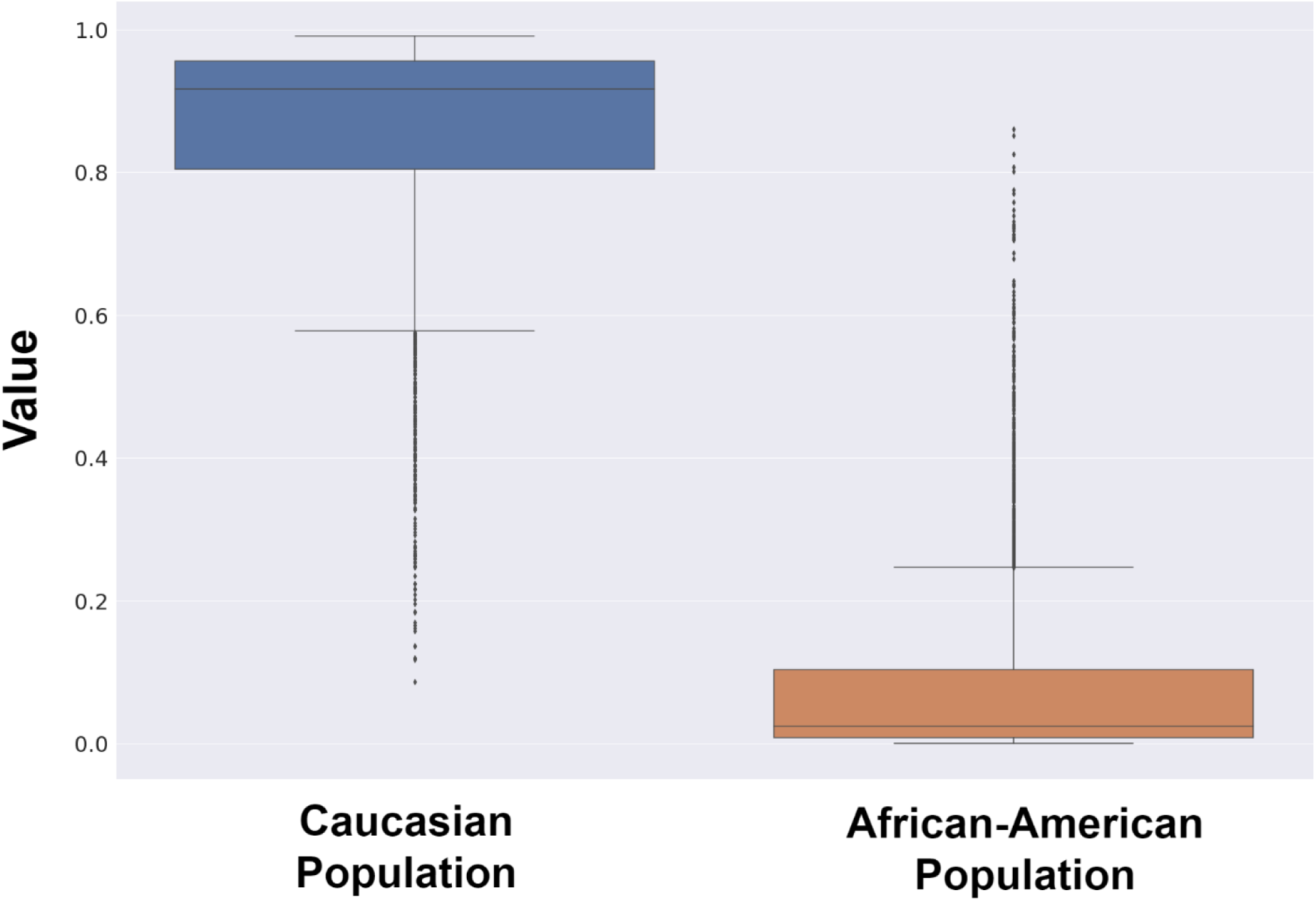
Box plots for African-American population percentage and for Caucasians’ percentage across counties.

As discussed also in the main paper, the Pearson correlation between obesity and diabetes prevalence is relatively high (0.698), they impacted the model differently. Figure 10 presents the box plots of *Diabetes* and *Obesity* features. Diabetes prevalence has low variance (5.76 * 10^−4^) between counties, leading to relatively random results when we compute the SHAP values. In contrast, the variance in obesity prevalence was much higher (2.02 * 10^−3^).

**Figure 9:**
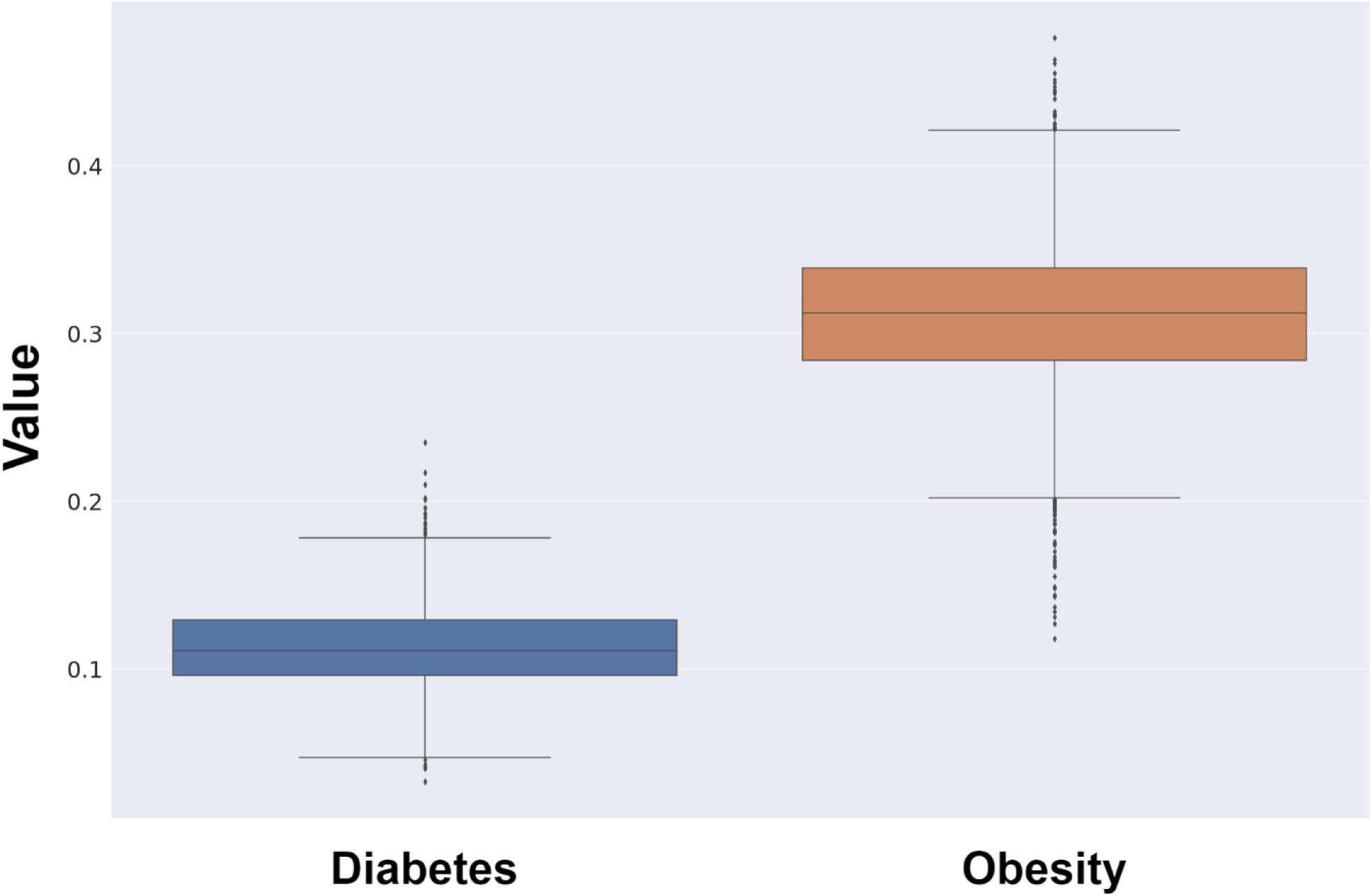
Box plots for Diabetes and Obesity percentage across counties.

## References

1. N. Itzhak, A. Nagori, E. Lior, M. Schvetz, R. Lodha, T. Sethi, and R. Moskovitch, “Acute hypertensive episodes prediction,” International Conference on Artificial Intelligence in Medicine (AIME), 2020.

2. T. Poteat, G. Millett, L. E. Nelson, and C. Beyrer, “Understanding covid-19 risks and vulnerabilities among black communities in america: The lethal force of syndemics,” Annals of Epidemiology, 2020.

3. M. W. Hooper, A. M. Napoles, and E. J. Pérez-Stable, “Covid-19 and racial/ethnic disparities,” Jama, 2020.

4. Y. Li, X. Cen, X. Cai, and H. Temkin-Greener, “Racial and ethnic disparities in covid-19 infections and deaths across us nursing homes,” Journal of the American Geriatrics Society, vol. 68, no. 11, pp. 2454–2461, 2020.

5. D. Fanelli and F. Piazza, “Analysis and forecast of covid-19 spreading in china, italy and france,” Chaos, Solitons & Fractals, vol. 134, p. 109761, 2020.

6. F. Carozzi, “Urban density and covid-19,” 2020.

7. Z. Sun, H. Zhang, Y. Yang, H. Wan, and Y. Wang, “Impacts of geographic factors and population density on the covid-19 spreading under the lockdown policies of china,” Science of The Total Environment, vol. 746, p. 141347, 2020.

8. A. Bhadra, A. Mukherjee, and K. Sarkar, “Impact of population density on covid-19 infected and mortality rate in india,” Modeling Earth Systems and Environment, pp. 1–7, 2020.

9. X. Wu, R. C. Nethery, B. M. Sabath, D. Braun, and F. Dominici, “Exposure to air pollution and covid-19 mortality in the united states,” medRxiv, 2020.

10. S. Chang, E. Pierson, P. W. Koh, J. Gerardin, B. Redbird, D. Grusky, and J. Leskovec, “Mobility network models of covid-19 explain inequities and inform reopening,” Nature, pp. 1–6, 2020.

11. G. Cahill, C. Kutac, and N. L. Rider, “Visualizing and assessing us county-level covid19 vulnerability,”

12. A. Tiwari, A. V. Dadhania, V. A. B. Ragunathrao, and E. R. Oliveira, “Using machine learning to develop a novel covid-19 vulnerability index (c19vi),” arXiv preprint 2009.10808, 2020.

13. G. A. Millett, A. T. Jones, D. Benkeser, S. Baral, L. Mercer, C. Beyrer, B. Honermann, E. Lankiewicz, L. Mena, J. S. Crowley, et al., “Assessing differential impacts of covid-19 on black communities,” Annals of Epidemiology, 2020.

14. A. Paul, P. Englert, and M. Varga, “Socio-economic disparities and covid-19 in the usa,” arXiv preprint 2009.04935, 2020.

15. C. A. Scannell, C. I. A. Oronce, and Y. Tsugawa, “Association between county-level racial and ethnic characteristics and covid-19 cases and deaths in the usa,” Journal of general internal medicine, vol. 35, no. 10, pp. 3126–3128, 2020.

16. S. M. Lundberg and S.-I. Lee, “A unified approach to interpreting model predictions,” in Advances in neural information processing systems, pp. 4765–4774, 2017.

17. A. Bishaw and K. G. Posey, “A comparison of rural and urban america: Household income and poverty,” Census blogs, pp. 855–902, 2016.

18. R.-U. C. Codes, “Usda economic research service. united states department of agriculture,” 2015.

19. E. A. Lundeen, S. Park, L. Pan, T. O’Toole, K. Matthews, and H. M. Blanck, “Obesity prevalence among adults living in metropolitan and nonmetropolitan counties—united states, 2016,” Morbidity and Mortality Weekly Report, vol. 67, no. 23, p. 653, 2018.

20. O. Ajilore and Z. Willingham, “Redefining rural america,” Center for American Progress, pp. 1–12, 2019.

## References

[1] Davis, J., Goadrich, M.: The relationship between precision-recall and roc curves. In: Proceedings of the 23rd international conference on Machine learning. pp. 233–240 (2006)

[2] Schulte, F., Lucas, E., Rau, J., Szabo, L., Hancock, J.: Millions of older americans live in counties with no icu beds as pandemic intensifies. Kaiser Health News (2020)

